# Estimating the impact of mobility patterns on COVID-19 infection rates in 11 European countries

**DOI:** 10.1101/2020.04.13.20063644

**Authors:** Patrick Bryant, Arne Elofsson

**Affiliations:** Science for Life Laboratory and Department of Biochemistry and Biophysics, Stockholm University, Stockholm, Sweden

**Author notes:** Corresponding Author: Patrick Bryant, Postal address: Box 1031, 171 62 Solna, Sweden.

## Abstract

**Background:** As governments across Europe have issued non-pharmaceutical interventions (NPIs) such as social distancing and school closing, the mobility patterns in these countries have changed. It is likely different countries and populations respond differently to the same NPIs and that these differences are reflected in the epidemic development.

**Methods:** We build a Bayesian model that estimates the number of deaths on a given day dependent on changes in the basic reproductive number, R_0_, due to changes in mobility patterns. We utilize mobility data from Google mobility reports using five different categories: retail and recreation, grocery and pharmacy, transit stations, workplace and residential. The importance of each mobility category for predicting changes in R_0_ is estimated through the model.

**Findings:** The changes in mobility have a large overlap with the introduction of governmental NPIs, highlighting the importance of government action for population behavioural change. The grocery and pharmacy sector is estimated to account for 97 % of the reduction in R_0_ (95% confidence interval [0·79,0·99]).

**Interpretation:** Our model predicts three-week epidemic forecasts, using real-time observations of changes in mobility patterns, which can provide governments with direct feedback on the effects of their NPIs. The model predicts the changes in a majority of the countries accurately but overestimates the impact of NPIs in Sweden and Denmark and underestimates them in France and Belgium.

**Funding:** Financial support: Swedish Research Council for Natural Science, grant No. VR-2016-06301 and Swedish E-science Research Center. Computational resources: Swedish National Infrastructure for Computing, grant No. SNIC-2019/3-319.

## Introduction

In December 2019 a new coronavirus (COVID-19) emerged in Wuhan, China. China implemented a quick strategy of suppression by locking the Wuhan province down on January 23^1^, and implementing social distancing procedures nationwide, with a successful outcome ^2^. Still, the virus rapidly spread across the world through our increasingly interconnected flight network, and shortly arrived in Europe. In February 2020 the number of cases started to increase rapidly in some European countries. To limit the spread of the virus, European countries introduced non-pharmaceutical interventions (NPIs) similar to China’s. These NPIs include social distancing, school closures, limiting international travel and lockdown^3^. All of these NPIs result in behavioural changes, which can be traced through mobility data from tracking the location of mobile phones.

Google recently released a time-limited sharing of mobility data^4^ from across the world as represented by summary statistics to combat COVID-19. The mobility data is measured in 6 different sectors: retail and recreation, grocery and pharmacy, parks, transit stations, workplace and residential. The effects of the government-issued NPIs are manifested through changes in these patterns, which are utilized in our model.

It is likely that different countries and populations respond differently to the same NPIs, why it is important to consider the effect of NPIs countrywise. By using real-life mobility data to model changes in the basic reproductive number, R_0_, the effects to NPIs across different countries can be modelled more accurately. The mobility data utilised here have some uncertainties and lack details but are the best openly available data source for tracking a population’s movement in all eleven studied countries. Governments can, in collaboration with telephone companies, obtain much more fine-grained data, enabling them to evaluate the effect of the NPIs in more detail.

After an initial rapid spread in China, control measures proved very successful to stop the spread both in China^5^ and in other parts of the world^6^,7. However, there is still a risk for subsequent spread upon lifting of these restrictions^7,8^. There is therefore an urgent need both for understanding and tracking the effects of governmental interventions and their removals. Large scale testing could provide valuable information about the effects of interventions, however, these are expensive, sometimes inaccurate and might violate privacy rights. In contrast, the use of large scale data from anonymous tracking of mobile phones is inexpensive and easily available.

Recently, a group from Imperial College released a report^7^ that estimates the effects of NPIs on R_0_. Their model is the basis for the model presented here. Their report had a large impact on how the UK government changed its intervention strategy^9^. A limitation of their model is the assumption that each intervention has the same impact in all countries, ignoring cultural and sociological differences. In contrast, by utilizing country specific mobility data in a Bayesian framework ^10^, we estimate the impact of each change in mobility pattern on R_0_. The resulting information provides an easy, straightforward way for governments to analyze if NPIs are working and to what extent. We show that in a three-week forecast our method provides a smaller mean error than the model from Imperial College.

## Methods

The model is trained on data from 30 days before the day after each country has observed 10 deaths in total up to (and including) 29 March, and then used to simulate a three week forecast from 30 March to 19 April.

### Model basis

Our model is based on the model used in the recent report^7^ from Imperial College London (ICL). The ICL report tries to estimate the impact of NPIs on the basic reproductive number (R_0_) in the same 11 countries modelled here. The main difference between the ICL model and the current one is the modelling of the impact on R_0_. The ICL team estimated the basic reproductive number at day t in country m (R_t,m_) as a function of the NPI indicators I_k,t,m_ in place at day t in country m as:

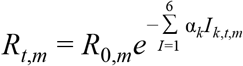

where I=1 when intervention k is implemented at day t in country m and α the impact of each intervention.

Here, we estimate R_t,m_ to be a function of the relative change in mobility pattern for each country:

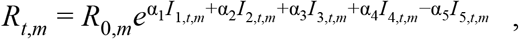

where I_1-5,t,m_ is the relative mobility in retail and recreation, grocery and pharmacy, transit stations, workplace and residential sectors respectively at day t in country m. The residential mobility parameter has a negative sign as an increase there is assumed to lower R_0_. We assume that the impact of each relative mobility change has the same relative impact across all countries and across time. Alpha is set to be gamma distributed with mean 0 5 and standard deviation 1. We did not include the data for the mobility category “Parks” as this data displayed much noise and cyclic peaks, as would be expected with varying weather^4^. The prior for R_0_ is set to:

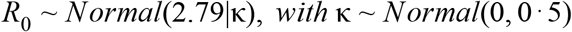

The value of 2.79 is chosen from the median value of a recent analysis of 12 modelling studies ^11^, and the normal distribution from ^2^.

The relative mobility is modelled as the relative value change compared to a mobility baseline estimated by Google^4^. The baseline is the median value, for the corresponding day of the week, during the 5-week period of 2020-01-03 to 2020-02-06. For the days for which no mobility data is available, the values were set to 0. The mobility data for the forecast (and days beyond the date for the last available mobility data) was set to the same values as the last observed days. The dates for the interventions were taken from the ICL report^7^, whose initial efforts were crowdsourced.

### Infection model

As the number of deaths in each country is likely to be the most accurate COVID-19 related data, we use this as the core of the model, being the posterior in the Bayesian simulations. The number of deaths in country m at day t is modelled as a negative binomial distribution with mean and variance accordingly:

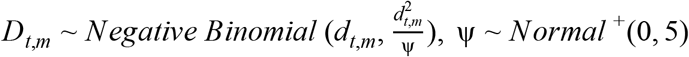

The expected number of deaths, d_t,m_, at day t in country m is given by:

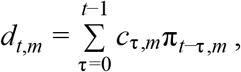

where π_*m*_ is the infection to death distribution in the country *m* given by a combination of the infection to onset distribution (Gamma(5.1,0·86)) and onset to death distribution (Gamma(18·8,0·45)) (combined with mean 23.9 days and standard deviation 0·45 days) times the infection fatality rate (*ifr*) ^7,12^:

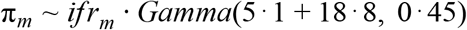

π_m_ is discretized in steps of 1 day accordingly:

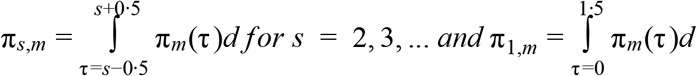

The *ifrs* are taken from previous estimates of the population at risk is about 1 %^13^ and adjusted for the predicted attack rate in the age group 50-59 years of age, assuming a uniform attack rate^7,8^,12, chosen due to having the least predicted underreporting in analyses of data from the Chinese epidemic ^12^. The number of deaths today is thus dependent on the cumulative number of cases from the previous days, weighted by the country-specific infection to death distribution.

The number of cases acquired at day τ in country m, *c*_τ,*m*_ is modelled with a discrete renewal process ^14,15^:

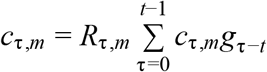, where *g*_τ−*t*_ ∼ *Gamma*(6·5, 0·62) (mean 6·5 days, standard deviation 0·62) is the serial interval distribution used to model the number of cases.

g_s_ is discretized in steps of 1 day accordingly:

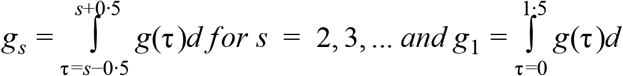

The number of cases today is thus dependent on the cumulative number of cases from the previous days, weighted by the serial interval distribution, times R_0_ at day t.

Just as in the ICL report^7^, we assume the starting point for the infection was 30 days before the day after each country has observed 10 deaths in total. From this assumed starting point, we initialize our model with 6 days ^2^ of cases drawn from an Exponential(0·03) distribution, which are inferred in the Bayesian posterior distribution (D_t,m_).

The implications on R_0_ due to relative mobility variations were estimated simultaneously for all countries in a hierarchical Bayesian framework using Markov-Chain Monte-Carlo (MCMC)^10^ simulations in Stan^16^. The death data^17^ used in the form of the number of deaths per day is from ECDC (European Centre of Disease Control), available and updated daily. We ran the model with eight chains, using 4000 iterations (2000 warm-up), as in the earlier work^7,16^. The parameter specifics of the simulation are available in the code (see below).

### MCMC Convergence

MCMC simulations are considered to converge when the Rhat statistics (a metric for comparing the variance between pooled and within-chain inferences) reach one^18^. A histogram of Rhat statistics for the modelled parameters in all simulation runs were constructed and analyzed. We also made sure no divergent transitions were observed by setting the adapt delta in the sampler (see code).

### Leave One Country Out Analysis

Since all countries are in different stages of their epidemics, different amounts of data are available for each country. To analyze how the model is influenced by different countries, we fit models using data from all countries except one using all 11 combinations^19^. We then estimate the importance of each mobility parameter in the leave-one-country-out analysis.

The relative difference in each mobility parameter provides an estimate of how each country affects R_0_ and thus the number of cases and deaths as well. Furthermore, the Pearson correlation coefficients for the mean R_0_ across all time points are calculated for each country in the different runs when all other 10 have been left out (see Figure S5).

### Forecast validation

To ensure the forecasts are reliable, we leave out three weeks of data (30 March - 19 April) and fit a model using data from the beginning of the epidemic up to the date for the beginning of the left-out data. We then evaluate the model with one week intervals from the 30th of March to the 19th of April. We evaluate by the average error and the average percent error (average error÷Σ observed deaths) during each of the three weeks, comparing with simulations obtained from the ICL model. We should note here that the ICL model does not converge for three-week predictions using 4000 iterations (see Figure S2).

### Code

All code is freely available at https://github.com/patrickbryant1/COVID19.github.io/ under the GPLv3 license.

## Results

### Estimating the cumulative number of cases, the number of deaths per day and changes in the basic reproductive number, R_0_

In Figure 1, for Italy and Sweden, and Figure S1, for all eleven modeled countries, our estimates of cumulative cases, daily deaths and the basic reproductive number R_0_ are shown. We simulate a three week forecast from 30 March to 19 April using data up to 29 March from the European Centre of Disease Control (ECDC) in the form of number of deaths per day, and relative mobility data estimated by Google^4^. According to the model, most countries appear to have their epidemic under control (April 19) (Table 1). The most successful country in terms of reducing R_0_ is Italy (R_0_≈0·19) and the least is Sweden (R_0_≈2·02).

**Table 1.** Mean estimates of R0 at the modelled start of the epidemic (when 10 cumulative deaths had been observed) and at the 29th of March for each respective country.

| Country | Modelled start of epidemic | Estimated mean $R_0$ at epidemic start | Estimated mean $R_0$ at 29 March | Relative change in Groceries and pharmacies on 29 March |
| --- | --- | --- | --- | --- |
| Austria | 2020-02-22 | 3.2 | 0.30 | -64% |
| Belgium | 2020-02-18 | 3.48 | 0.46 | -53% |
| Denmark | 2020-02-21 | 3.19 | 1.30 | -22% |
| France | 2020-02-07 | 3.15 | 0.27 | -72% |
| Germany | 2020-02-15 | 3.34 | 0.52 | -51% |
| Italy | 2020-01-27 | 3.54 | 0.19 | -85% |
| Norway | 2020-02-24 | 2.82 | 0.81 | -32% |
| Spain | 2020-02-09 | 3.53 | 0.26 | -76% |
| Sweden | 2020-02-18 | 3.04 | 2.02 | -10% |
| Switzerland | 2020-02-14 | 2.97 | 0.47 | -51% |
| United Kingdom | 2020-02-12 | 2.87 | 0.53 | -46% |

**Figure 1.**
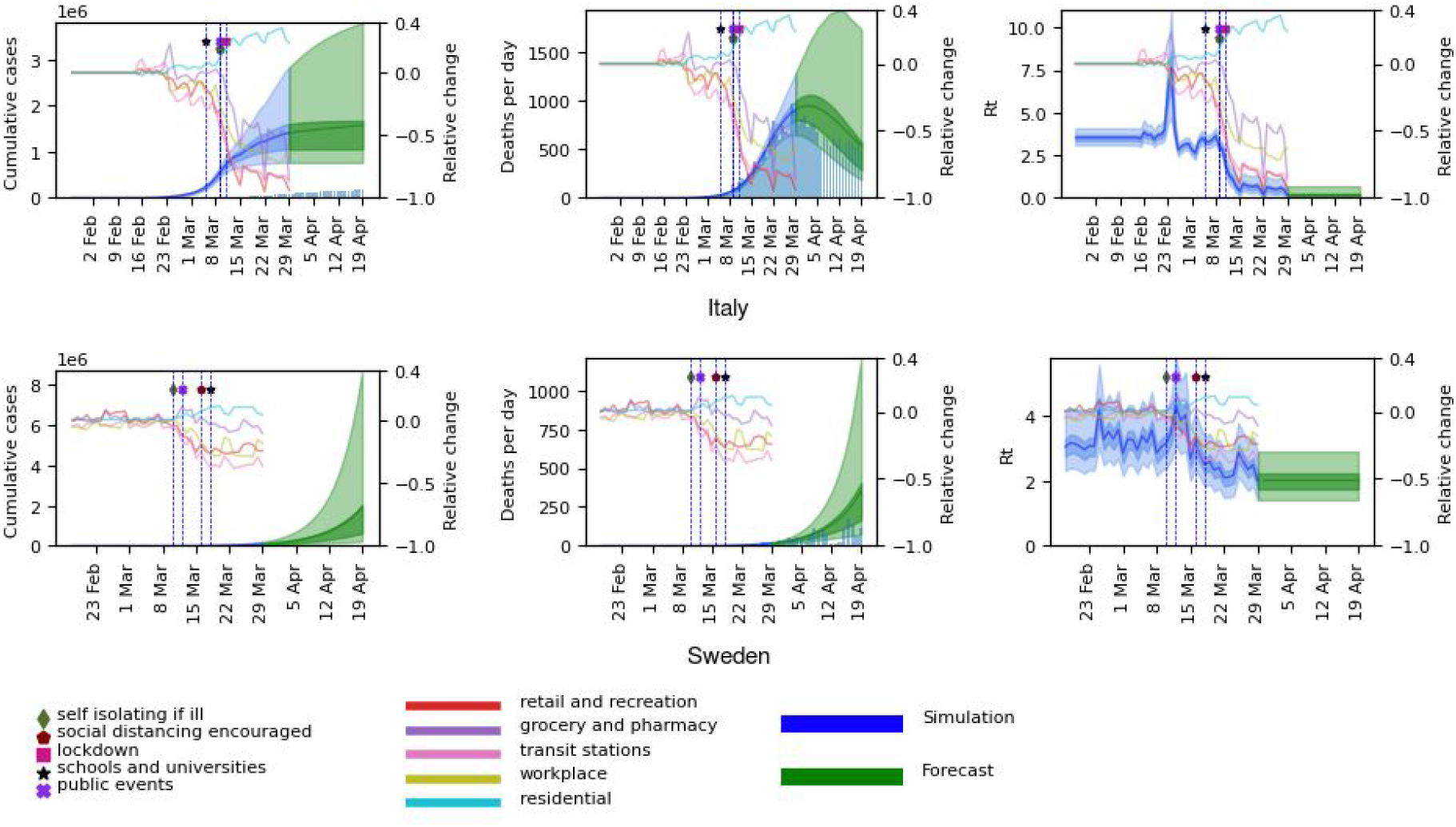
Model results in the form of cumulative number of cases, deaths per day and R_0_ for Italy and Sweden, are displayed on the left axes. The model results start from 30 days before 10 accumulated deaths had been observed. The blue curves represent the estimations so far, while the green represents a three week forecast (30 March-19 April). The 50 % and 95 % confidence intervals are displayed in darker and lighter shades respectively, with the mean as a solid line. The histograms represent the number of cases and deaths reported by the European Center for Disease Control (ECDC). Mobility data for the five modelled sectors represented in terms of relative change compared to baseline (observed in a five-week period of 2020-01-03 to 2020-02-06) is displayed on the right axes. The dates for the introduction of different NPIs are marked with vertical lines. As can be seen, the NPIs have very strong implications for the mobility patterns. The mobility data ranges from 2020-02-15 to 2020-03-29, after which the final levels are fixed.

From Figure S1, it can be seen that in all countries the interventions have some positive effect, decreasing the estimated R_0_ between the epidemic start and March 29. It can be noted that during the development of the epidemic, R_0_ displays a wide range of values. In some countries, the mean of the estimated R_0_ displays a rapid increase to values as high as 15, coupled with an increase in mobility (primarily) to grocery and pharmacies exactly when the interventions are put into force (see Figures 1 and S1).

The estimated number of deaths for up to three weeks after the model is trained, have a good correspondence with the observed number (Figures 1, S1 and Table 2). Compared with the Imperial College London (ICL) model^7^, our model displays both lower errors and less uncertainty (see Figures 2, S2 and Table S1). The average absolute errors over the 11 countries in the number of deaths are lower across all three weeks (week 1: 68 vs 158, week 2: 119 vs 488, and week 3: 113 vs 1497 for ours and the ICL-teams respectively).

**Table 2.** Average error and average fractional error in the number of deaths for each country between the mean predicted number of deaths per day and the observed number in one, two and three week forecasts respectively. A corresponding table for the ICL model can be found in Table S2.

| Three week predictions for the number of deaths per day |  |  |  |  |  |  |
| --- | --- | --- | --- | --- | --- | --- |
|  | Average error |  |  | Average percent error |  |  |
| Country | week 1 | week 2 | week 3 | week 1 | week 2 | Week 3 |
| Austria | -3 | -7 | -2 | -2.6 | -4.4 | -2.1 |
| Belgium | -46 | -179 | -183 | -5.0 | -8.7 | -8.7 |
| Denmark | 0 | 8 | 24 | -0.2 | 8.4 | 28.4 |
| France | -310 | -424 | -395 | -5.9 | -6.8 | -7.2 |
| Germany | -24 | -10 | -22 | -2.5 | -0.7 | -1.4 |
| Italy | 177 | 254 | 94 | 3.3 | 6.2 | 2.5 |
| Norway | 0 | 0 | 2 | 0.8 | 0.4 | 3.9 |
| Spain | -68 | 142 | 132 | -1.1 | 3.1 | 3.6 |
| Sweden | -5 | 19 | 151 | -1.9 | 3.7 | 24.2 |
| Switzerland | 11 | 37 | 45 | 3.6 | 12.9 | 16.2 |
| United Kingdom | -103 | -231 | -191 | -3.1 | -4.2 | -3.4 |
| Average absolute average | 68 | 119 | 113 | 2.7 | 5.4 | 9.2 |

**Figure 2.**
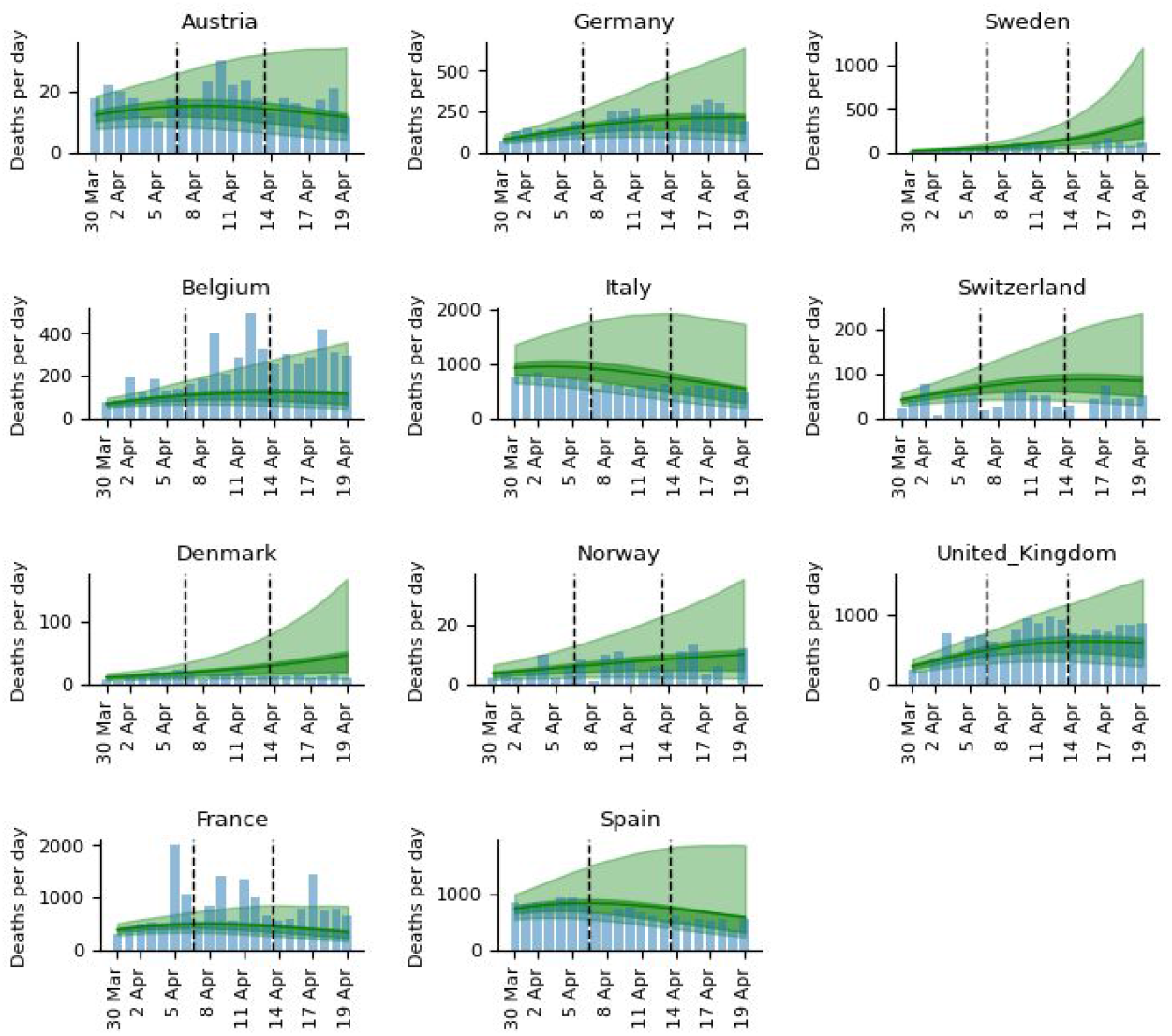
Three week predictions for all countries in the form of deaths per day for the weeks 1: (Mar 30 - April 5), week 2 (April 6 - April 12) and week 3 (April 13 - April 19). The 50 % and 95 % confidence intervals are displayed in darker and lighter shades respectively, with the mean as a solid line. The blue histogram represents the observed deaths.

### Comparing mobility data across countries

When overlaying the implementation dates of the NPIs with the mobility data, it is clear that governmental decisions have a very large impact on the populations in the 11 modelled countries (see Figure S1). Most countries display very similar relative changes in their mobility patterns, with mobility in retail and recreation, grocery and pharmacy, transit stations and workplace decreasing and mobility in the residential category increasing.

Most countries have similar relative changes across the sectors (Figure S1). The ones that display smaller relative changes (Denmark, Norway and Sweden) also display smaller reductions in R_0_, which is a natural consequence of our model, as it assumes that changes in R_0_ are directly related to changes in mobility. The mobility patterns in Sweden display barely half of the relative changes compared with France, Spain, and Italy, and the reduction in R_0_ is therefore smaller in Sweden.

### The importance of mobility sectors for modelling changes in R_0_

Analyzing the importance of each mobility parameter for predicting the reduction in R_0_ shows that the grocery and pharmacy sector appears to be the clearest indicator for R_0_ change (see Figure 3). The grocery and pharmacy parameter is estimated to account for 97 % of the reduction in R_0_ with a narrow confidence interval (CI). The residential parameter seems important as well, which would be expected, but the confidence interval displays a large uncertainty.

**Figure 3.**
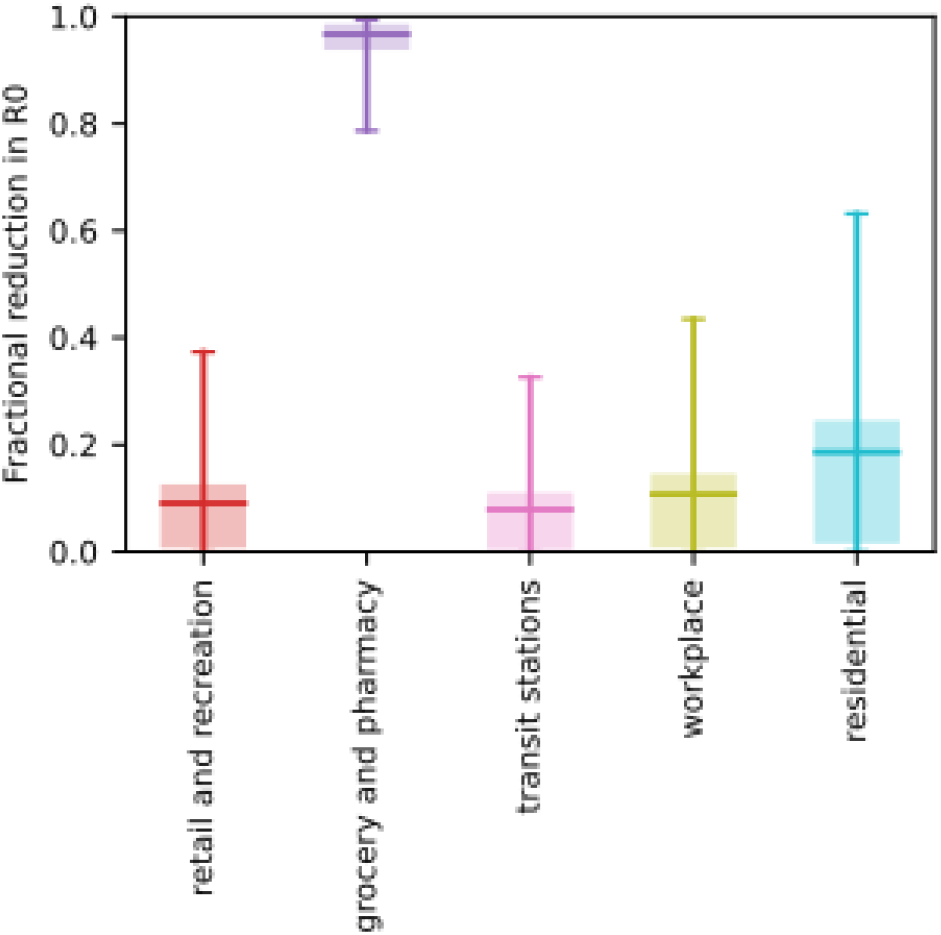
Estimation of the importance of each mobility parameter for predicting the reduction in R_0_. The five different modelled sectors are shown with marked means, 50 % (boxes) and 95 % confidence intervals (end points). The grocery and pharmacy sector appears to be the clearest indicator for R_0_ change, estimated to account for 97 % (95% confidence interval [0·79,0·99]) of the reduction in R_0_. The residential parameter seems important as well, which would be expected, but the CI displays a large uncertainty.

### Model validation

The means and CIs for the mobility parameters (see Figure S4) are almost identical in the leave-one-country-out analysis (LOO) analysis. A very wide CI is observed for Italy in the grocery and pharmacy sector though, emphasizing the importance of the Italian data. The variable R_0_ values in the LOO analysis show high Pearson correlations, with Italy and especially the United Kingdom displaying lower correlations (see Figure S5). Italy and the United Kingdom correlate quite badly with each other. One of 4000 iterations ended with a divergence (0·025 %) when France and Spain were excluded. A histogram of Rhat statistics for the modelled parameters in all simulations for the main analysis is displayed in Figure S6.

## Discussion

Our model makes it clear that the non-pharmaceutical interventions (NPIs) introduced by governments across Europe have had substantial effects on both mobility patterns and in preventing the spread of COVID-19. By tracking the relative change in mobility in the grocery and pharmacy sector it is possible to account for 97 % of the reduction in the basic reproductive number, R_0_, in our model. This information provides an easy, straightforward way for governments to analyze if NPIs are working and to what extent.

Why the grocery and pharmacy sector has the biggest impact on estimated changes in R_o_ is not clear. It is possible that this sector enables contacts between different communities, but this requires further analysis to be fully understood. Since R_0_ is strongly dependent on the changes in mobility, rapid changes in mobility leads to rapid changes in R_0_. This has drastic consequences to the estimated development of the epidemic in a country.

However, changes in R_0_ will not manifest in the number of deaths per day until about three weeks later (the mean value in the gamma distribution for infection to death is 23·9 days, see methods section). Therefore, we provide a three-week forecast. The estimates have a good correspondence with the observed numbers in most countries (see Figures 2 and Table 2), and compared with the ICL-model, our model displays both lower errors and less uncertainty (Figures 2, S2 and Tables 2,S1). It can also be noted that the ICL model overpredicts the number of deaths in all countries at the end of the estimate.

The estimated number of cases has great uncertainty across all countries. It should be noted here that one limitation of our model is that it does not take herd-immunity effects into account, which should be reached when around 60-80 % of the population is infected ^20^, but it is unlikely that sufficiently high infection has been reached yet for this to have a significant effect. Another limitation of the model is the assumption that the impact of each relative mobility change has the same relative impact across all countries and across time. Likely both more detailed mobility data and intermixing patterns need to be considered, metrics that are not available.

The number of cases are also highly dependent on having the correct infection-fatality-rate (*ifr*). This quantity is only modelled for the age group 50-59 years and does thereby not take into account the attack rates for the whole of each country’s population (see methods section). If a country managed to avoid the elderly being infected, that would lower the *ifr* ^*21*^, which could explain prediction differences to some extent.

The model validation, both by a leave-one-country-out analysis and by predicting a three week forecast, ensures the model’s robustness. The countries where the errors stand out are Denmark and Sweden, with over-predicted estimates, and Belgium and France, under-predicted. We note that these two pairs of countries are close both geographically and culturally^22,23^, possibly explaining the systematic differences. The differences may also be caused by differences in reporting between the countries^24,25^. For instance, on April 5 more than 2000 deaths were reported in France, due to sudden inclusion of potential COVID-19 attributed deaths in nursing homes occuring at earlier dates ^26^.

Here, we present a model to estimate the effects of public interventions on the spread of COVID-19 that does not assume that interventions have identical effects in different geographical and cultural settings. In contrast, our model uses *observational* data of mobility patterns in five environments to estimate changes in the transmission rate. Our model creates the possibility to track rapid changes in spread, right now and predict their consequences three weeks ahead in time. This enables governments to use anonymous real-time data to adjust their policies. We do foresee that such models will become incrementally more powerful as more detailed mobility data becomes available in the future.

## Data Availability

All code is freely available at https://github.com/patrickbryant1/COVID19.github.io/ under the GPLv3 license.
Data and future predictions will be made available at https://covid19.bioinfo.se/

https://github.com/patrickbryant1/COVID19.github.io/

https://covid19.bioinfo.se/

## Contributors

PB designed and implemented the study. PB generated all figures and wrote the initial draft of the manuscript, which was further edited, reviewed, revised and approved by both authors. AE provided the initial extraction of the mobility data (before it was made available in .csv format).

## Availability

Data and future predictions will be made available at https://covid19.bioinfo.se/

## Acknowledgements

We acknowledge Claudio Bassot’s contribution by sharing both the Imperial College London report and the Google mobility data. Without this information, this study would not be possible. We are also grateful to various colleagues and friends that contributed to the discussion. Finally we thank the authors of the Imperial College London report to make their data and model freely available.

## Supplementary Material

### Figures

**Figure S1.**
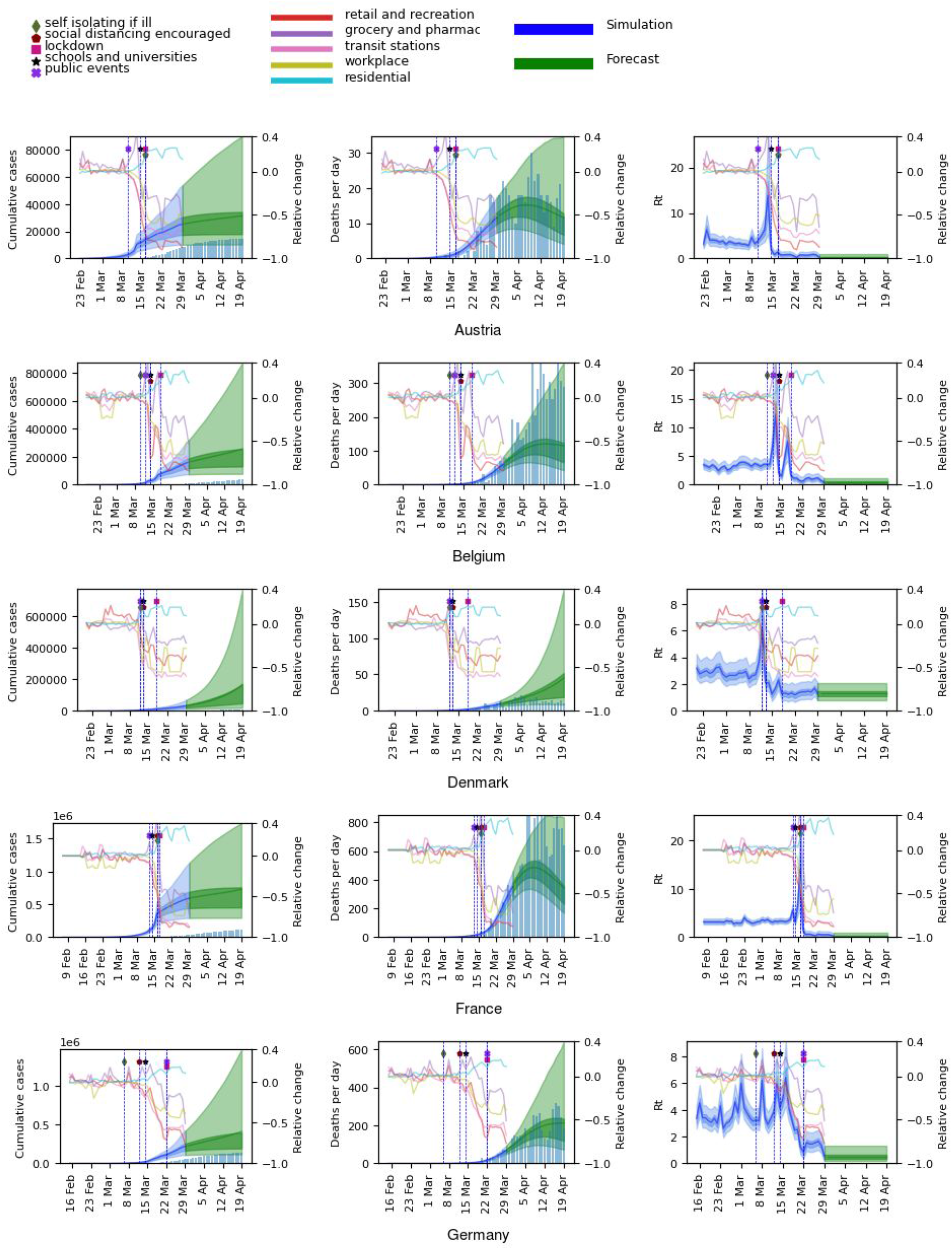
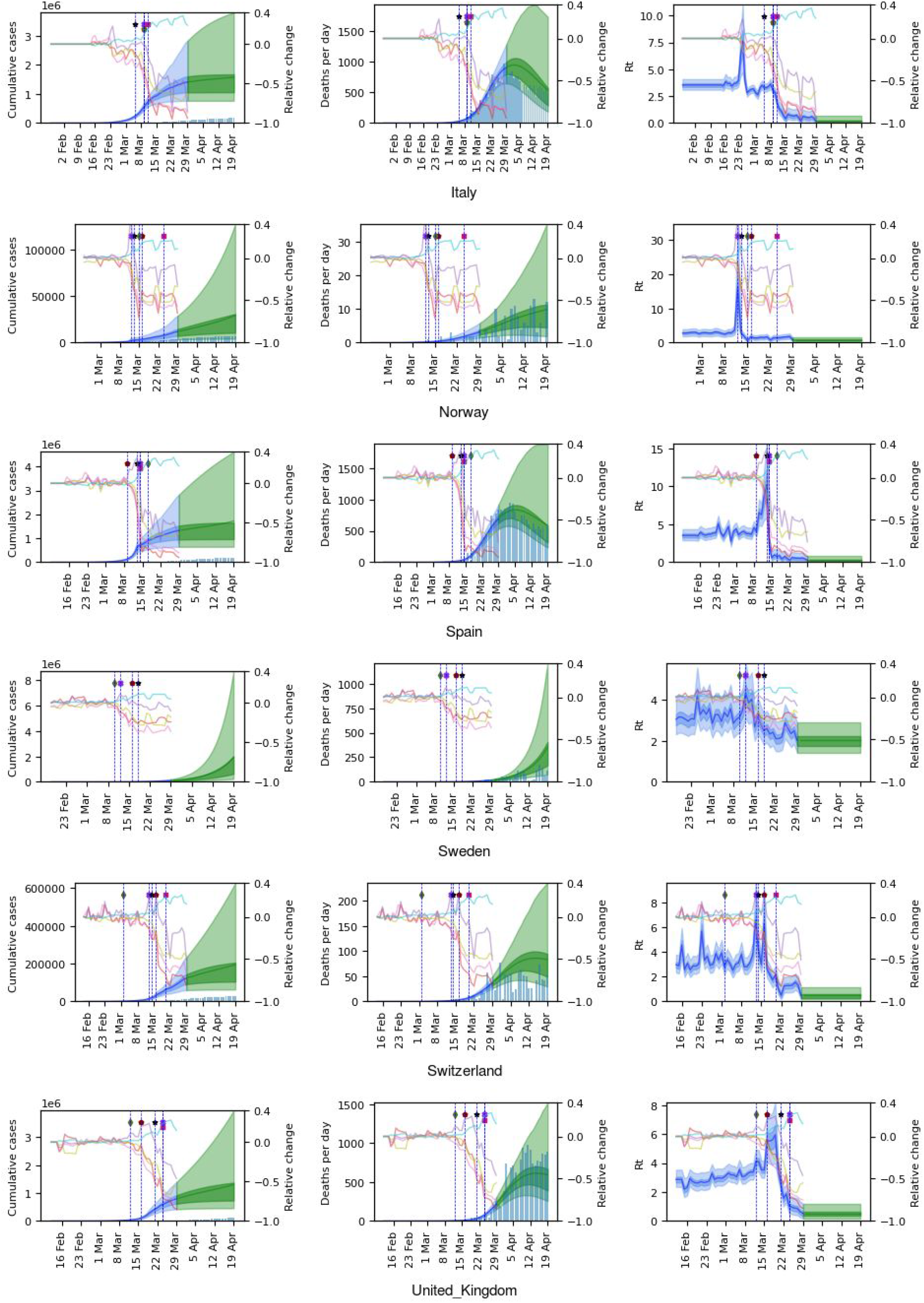
Model results in the form of cumulative number of cases, deaths per day and R_0_ for each respective country, are displayed on the left axes. The model results start from 30 days before 10 accumulated deaths had been observed. The blue curves represent the estimations so far, while the green represents a three week forecast (30 March-19 April). The 50 % and 95 % confidence intervals are displayed in darker and lighter shades respectively, with the mean as a solid line. The histograms represent the number of cases and deaths reported by the European Center for Disease Control (ECDC). Mobility data for the five modelled sectors represented in terms of relative change compared to baseline (observed in a five-week period of 2020-01-03 to 2020-02-06) is displayed on the right axes. The dates for the introduction of different NPIs are marked with vertical lines. As can be seen, the NPIs have very strong implications for the mobility patterns. The mobility data ranges from 2020-02-15 to 2020-03-29, after which the final levels are fixed.

**Figure S2.**
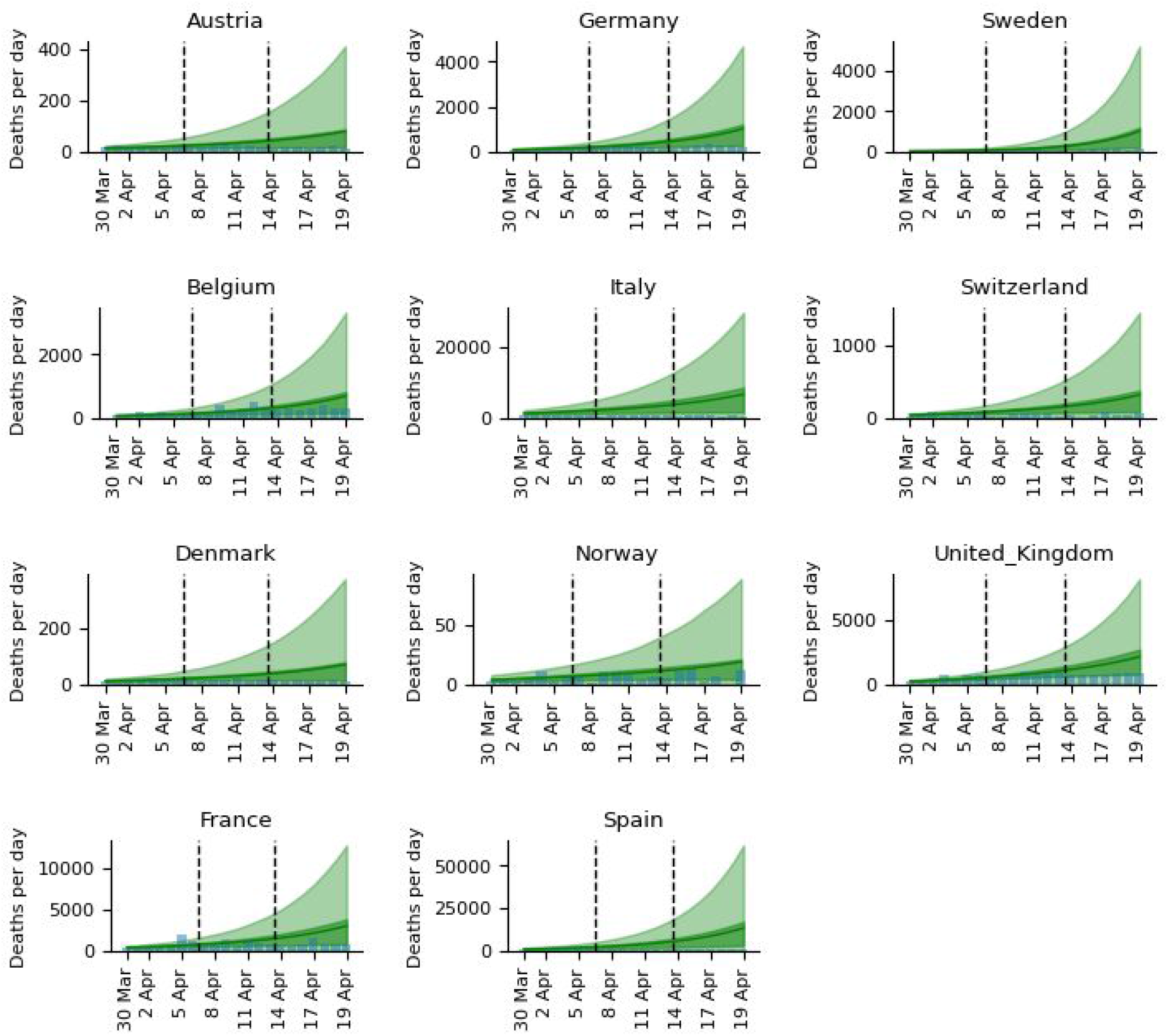
ICL model. Three week predictions for all countries in the form of deaths per day for the weeks Mar 30 - April 5, April 6 - April 12 and April 13 - April 19. The 50 % and 95 % confidence intervals are displayed in darker and lighter shades respectively, with the mean as a solid line. The blue histogram represents the observed values.

**Figure S3.**
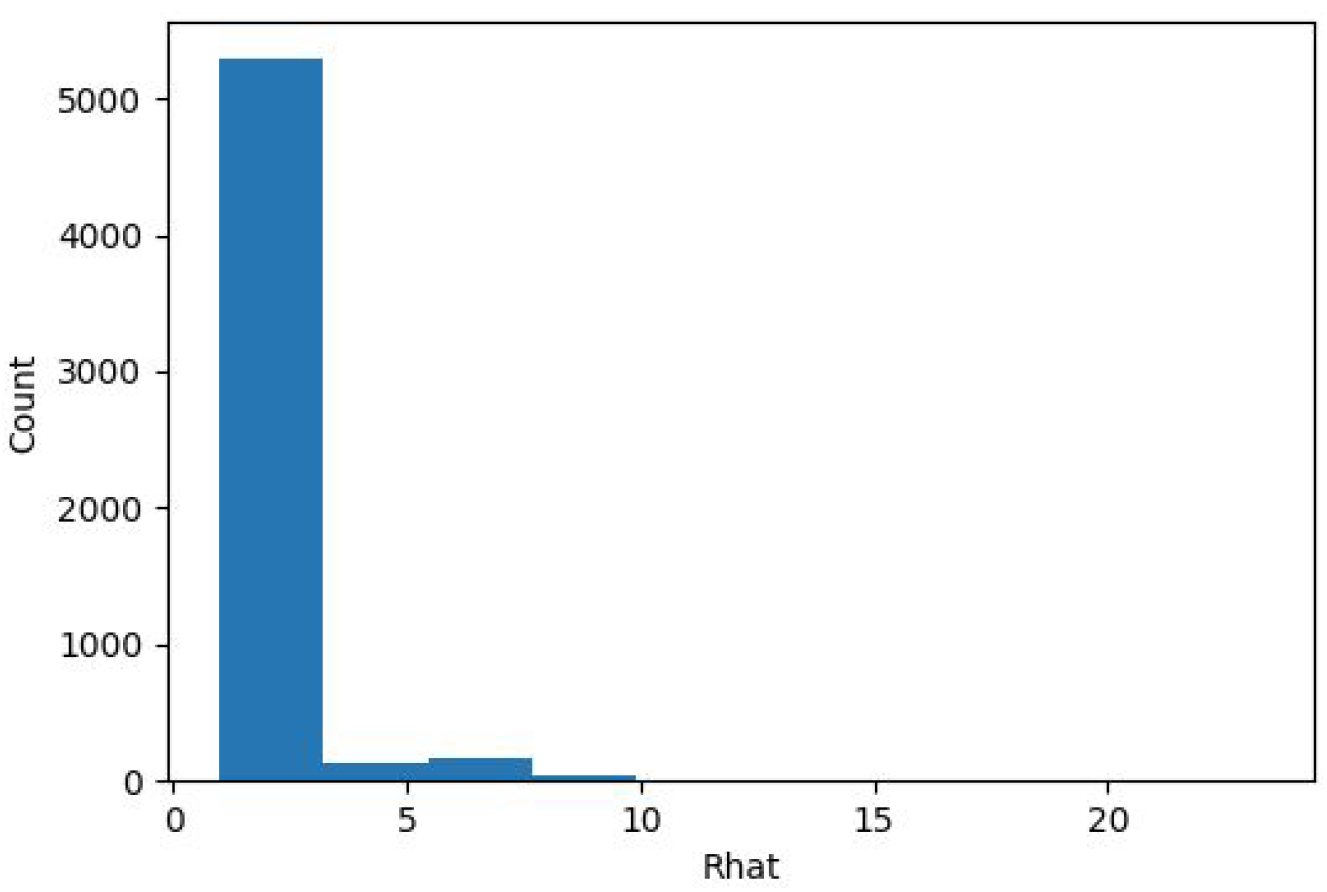
Rhat statistics for all simulation parameters in the Imperial College London model (Flaxman, Mishra, Gandy et al. 2020) using all 11 countries. Values of 1 indicate convergence in the simulations.

**Figure S4.**
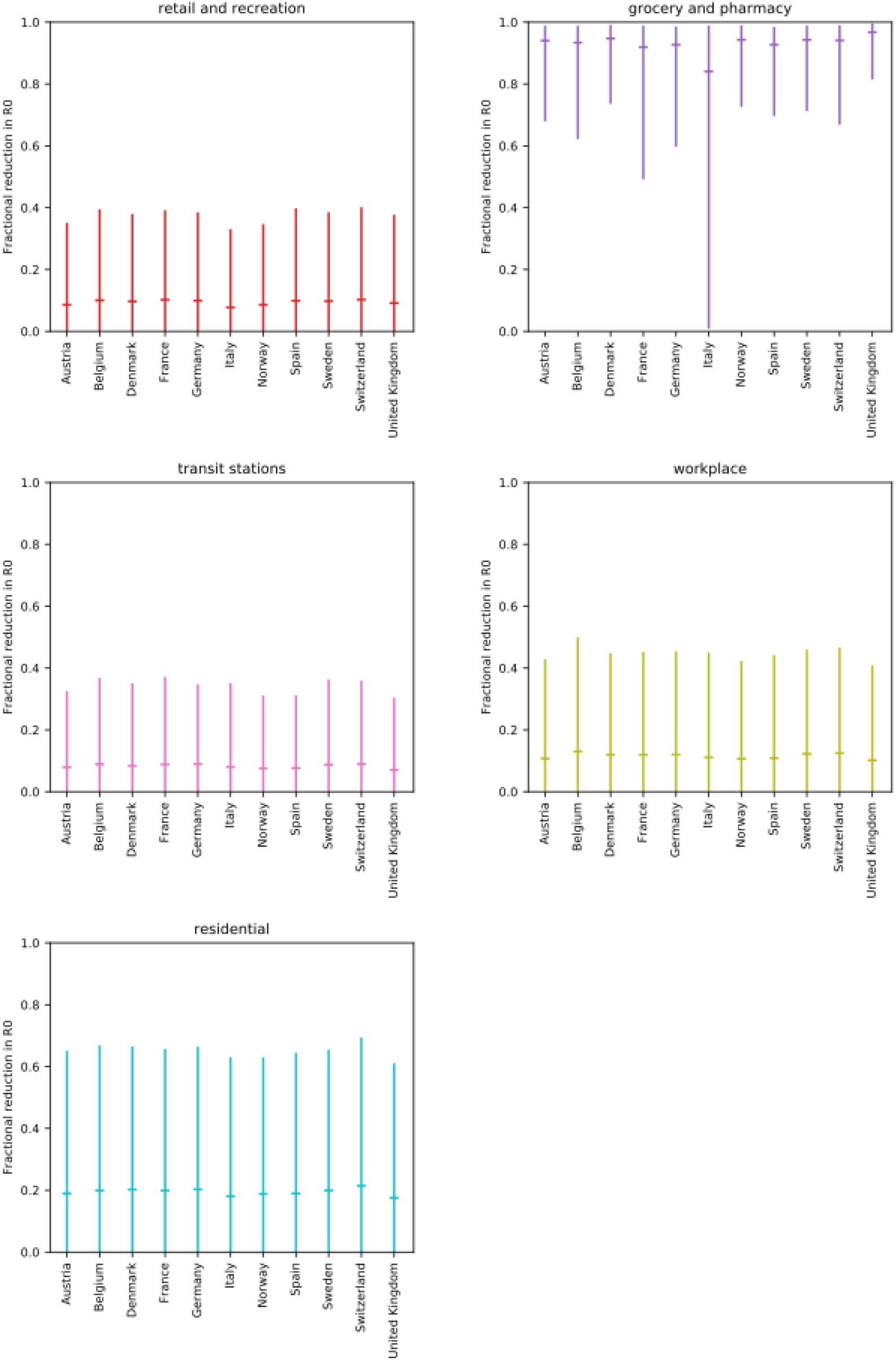
Estimation of the importance of each mobility parameter for predicting the reduction in R_0_ from the leave-one-country-out analysis. The x-axis indicates which country has been left out in the simulation. The five different modelled sectors are shown with marked means and 95 % confidence intervals (CIs). As can be seen, the means and CIs are very similar, regardless of which country that has been left out.

**Figure S5.**
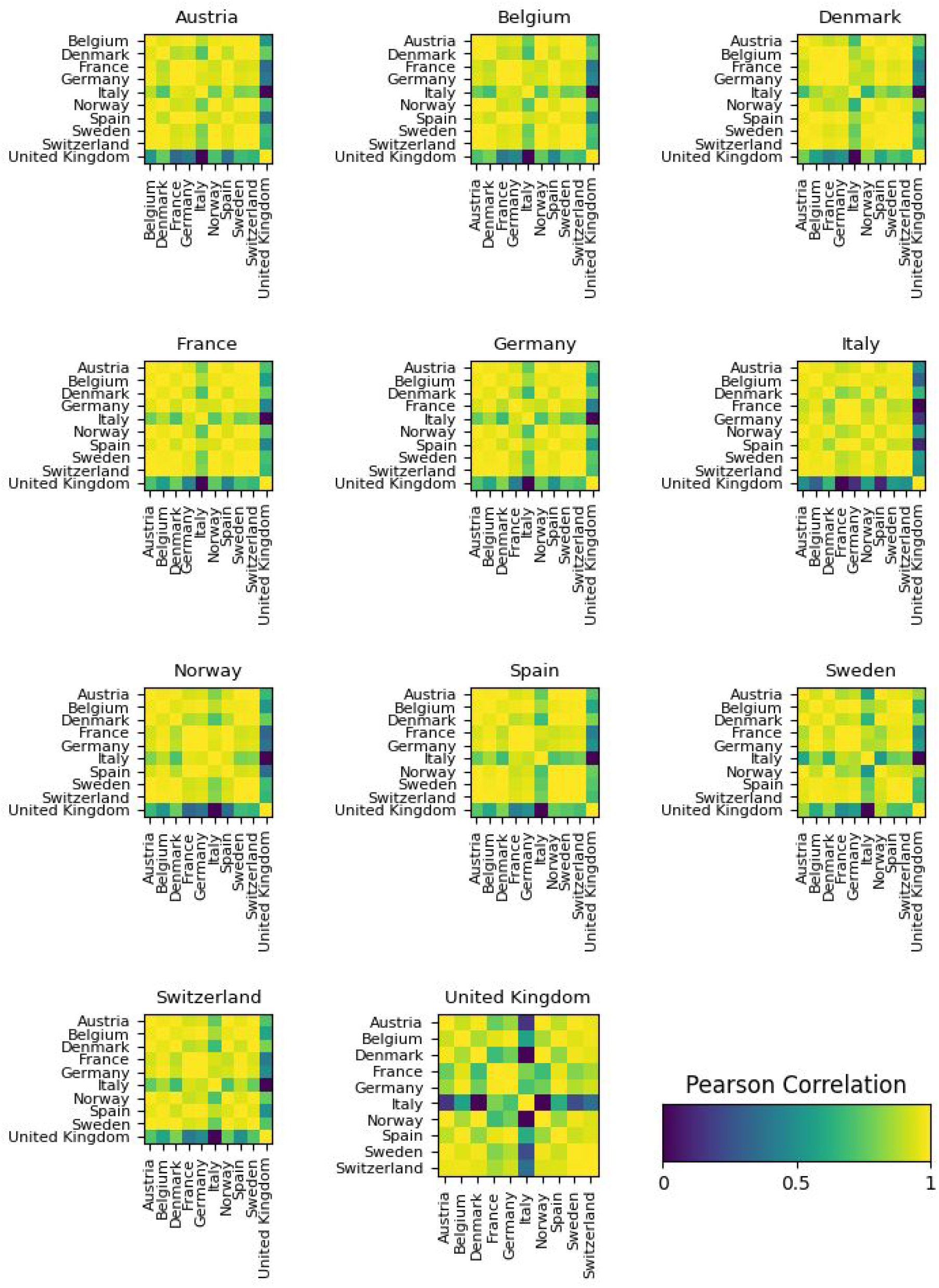
Visualization of the Pearson correlation coefficients for the mean R_0_ across all timepoints (including the forecast) for each country in the different runs when all other 10 (one per run) have been left out. Italy creates 2 clusters of countries that seem to fit together. Austria and the United Kingdom do not seem to be influenced much by any of the other countries.

**Figure S6.**
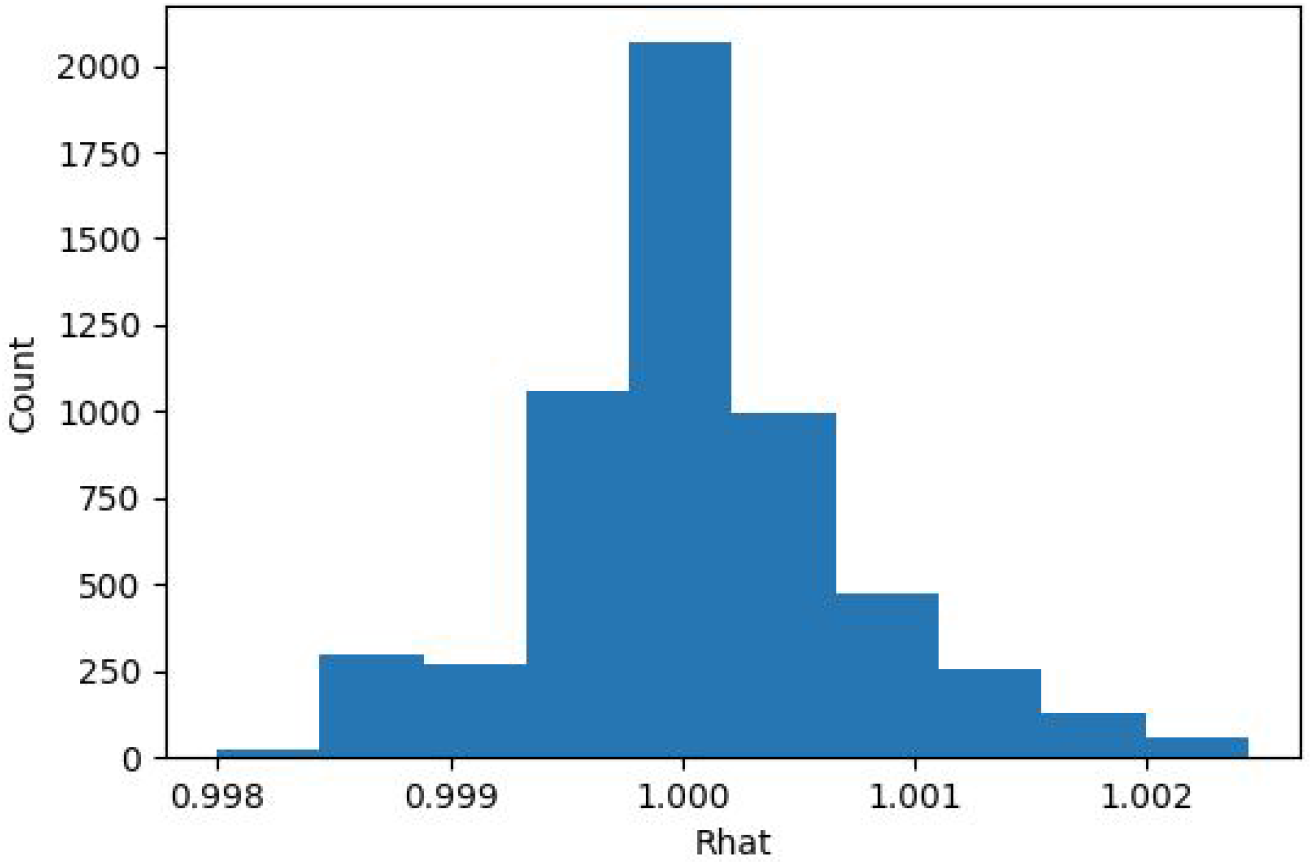
Rhat statistics for all simulation parameters using all 11 countries. Values of 1 indicate convergence in the simulations.

### Tables

**Table S1.** Average error and average fractional error in the number of deaths for each country between the mean predicted number of deaths per day in a one week forecast and the observed number. Results for both our model (Mobility model) and the model from the Imperial College London team (ICL model) are shown.

| Comparison of three week predictions between the Mobility and the ICL model |  |  |  |  |  |  |  |  |  |  |  |  |
| --- | --- | --- | --- | --- | --- | --- | --- | --- | --- | --- | --- | --- |
|  | Error in predicted deaths |  |  |  |  |  | Average percent error |  |  |  |  |  |
|  | Mobility | ICL | Mobility | ICL | Mobility | ICL | Mobility | ICL | Mobility | ICL | Mobility | ICL |
| Country | 30 Mar - 5 Apr | 30 Mar - 5 Apr | 6 Apr - 12 Apr | 6 Apr - 12 Apr | 13 Apr - 19 Apr | 13 Apr - 19 Apr | 30 Mar - 5 Apr | 30 Mar - 5 Apr | 6 Apr - 12 Apr | 6 Apr - 12 Apr | 13 Apr - 19 Apr | 13 Apr - 19 Apr |
| Austria | -3 | 0 | -7 | 9 | -2 | 43 | -2.6 | 0.0 | -4.4 | 5.6 | -2.1 | 40.7 |
| Belgium | -46 | -41 | -179 | -93 | -183 | 175 | -5.0 | -4.4 | -8.7 | -4.5 | -8.7 | 8.3 |
| Denmark | 0 | 1 | 8 | 13 | 24 | 40 | -0.2 | 1.0 | 8.4 | 12.8 | 28.4 | 46.6 |
| France | -310 | -245 | -424 | 130 | -395 | 1390 | -5.9 | -4.7 | -6.8 | 2.1 | -7.2 | 25.3 |
| Germany | -24 | -16 | -10 | 97 | -22 | 464 | -2.5 | -1.7 | -0.7 | 7.3 | -1.4 | 28.6 |
| Italy | 177 | 829 | 254 | 2175 | 94 | 4528 | 3.3 | 15.5 | 6.2 | 52.9 | 2.5 | 120.5 |
| Norway | 0 | 1 | 0 | 2 | 2 | 8 | 0.8 | 1.9 | 0.4 | 3.5 | 3.9 | 15.3 |
| Spain | -68 | 462 | 142 | 2682 | 132 | 8367 | -1.1 | 7.6 | 3.1 | 58.2 | 3.6 | 226.7 |
| Sweden | -5 | -2 | 19 | 64 | 151 | 490 | -1.9 | -0.8 | 3.7 | 12.5 | 24.2 | 78.5 |
| Switzerland | 11 | 11 | 37 | 69 | 45 | 190 | 3.6 | 3.5 | 12.9 | 23.6 | 16.2 | 68.1 |
| United Kingdom | -103 | -134 | -231 | -36 | -191 | 776 | -3.1 | -4.1 | -4.2 | -0.6 | -3.4 | 13.9 |
| Total absolute average | 68 | 158 | 119 | 488 | 113 | 1497 | 2.7 | 4.1 | 5.4 | 16.7 | 9.2 | 61.1 |

